# Polyautoimmunity Clusters as a New Taxonomy of Autoimmune Diseases

**DOI:** 10.1101/2021.08.15.21262029

**Authors:** Manuel Rojas, Carolina Ramírez-Santana, Yeny Acosta-Ampudia, Diana M. Monsalve, Mónica Rodriguez-Jimenez, Elizabeth Zapata, Angie Naranjo-Pulido, Ana Suárez-Avellaneda, Lady J. Ríos-Serna, Carolina Prieto, William Zambrano-Romero, María Alejandra Valero, Rubén D. Mantilla, Chengsong Zhu, Quan-Zhen Li, Carlos Enrique Toro-Gutiérrez, Gabriel J. Tobón, Juan-Manuel Anaya

**Affiliations:** Center for Autoimmune Diseases Research (CREA), School of Medicine and Health Sciences, Universidad del Rosario; Bogota, Colombia; Centro de Referencia en Osteoporosis, Reumatología & Dermatología; Cali, Colombia; Centro de Investigación en Reumatología, Autoinmunidad y Medicina Traslacional (CIRAT), Universidad ICESI, Cali; Colombia; Dermatology and Rheumatology Foundation (FUNINDERMA); Bogota, Colombia; Department of Immunology, Microarray & Immune Phenotyping Core Facility, University of Texas Southwestern Medical Center; Dallas, USA; Clinica del Occidente; Bogota, Colombia

## Abstract

Polyautoimmunity (PolyA) is an emerging concept that may help to develop a better classification of autoimmune diseases (ADs). Thus, we aimed to develop new taxonomy based on PolyA. Two-hundred and fifty-four consecutive patients were included with rheumatoid arthritis (RA, n:146), systemic lupus erythematosus (SLE, n:45), Sjögren’s syndrome (SS, n:29), autoimmune thyroid disease (AITD, n:17) and systemic sclerosis (SSc, n:17). Clinical features, autoantigen array chip, lymphocytes immunophenotype and cytokine profile were assessed simultaneously. The coexistence of two or more ADs with classification criteria was termed *“Overt PolyA”*, whereas the presence of autoantibodies unrelated to the index AD, without criteria fulfillment, was named *“Latent PolyA”*. Combination of IgG autoantibodies yielded high accuracy for classification of ADs. In SLE, Histone H2A, Sm/RNP, ssDNA, and dsDNA IgG autoantibodies were the most predictive autoantibodies for this condition. Laminin, Ro/SSA (52 kDa), and U1−snRNP B/B’ for SS; Thyroglobulin for AITD; Ribo Phosphoprotein P1, and CENP-A for SSc. Interestingly, Thyroglobulin and U1−snRNP B/B’ were mutual diagnostic biomarkers in SS and SSc. Latent PolyA showed in nearly 70% of patients, whereas overt PolyA was most common in AITD (82.4%) and SLE (40%). Cluster analysis based on autoantibodies yielded three clusters of which clusters 2 and 3 exhibited high frequency of latent and overt PolyA with distinctive clinical and immunological phenotypes. Combination of autoantibodies demonstrated high performance for classification of ADs. Patients with both latent and overt PolyA cluster together and exhibit differential clinical and immunological features. High prevalence of latent and overt PolyA advocates for routinary surveillance in clinical settings.

**One Sentence Summary:** This is a proof-of-concept study which allowed a new classification of autoimmune diseases. The results highlight that most patients with both latent and overt polyautoimmunity cluster together, with differential clinical and immunological characteristics.

## INTRODUCTION

Autoimmune diseases (ADs) may manifest as a single AD, or concurrently with other ADs, a condition named polyautoimmunity (PolyA) *(1)*. In the same spectrum, the coexistence of two or more ADs with classification criteria is termed “Overt PolyA”, whereas the presence of autoantibodies unrelated to the index AD, without criteria fulfillment, is named “Latent PolyA” *(2)*. In addition, both conditions can coexist in a single patient *(3)*. Although, the clinical and immunological relevance of this classification is still unknown, it is established that recognition of differential immunological patterns on autoimmunity may allow the implementation of personalized strategies in the management of ADs *(4)*.

Recent studies in systemic lupus erythematosus (SLE) showed that autoantibody-based classification allowed the identification of subgroups associated with disease activity, and inflammatory cytokines *(5)*. In addition, another study showed the association between IL-12/23p40 and overt PolyA *(3)*. Thus, suggesting a differential expression of cytokines, that can be used as therapeutic targets in patients with PolyA.

It is well recognized that the development of autoantibodies precedes clinical manifestations of ADs *(6)*, and combinations of autoantibodies are predictive for disease evolution *(7)*. Thus, it is likely that patients with latent PolyA could develop overt PolyA in the future *(2, 3)*. In this line, studies on rheumatoid arthritis (RA) showed that IgG or IgA citrullinated peptide antibodies and IgA rheumatoid factor precede the appearance of RA by several years *(8–10)*. This was similar for SLE *(11)*, autoimmune thyroid disease (AITD) *(12–14)*, systemic sclerosis (SSc) *(15, 16)*, and Sjögren’s syndrome (SS) *(17)*.

Family-based studies showed that first-degree relatives of patients with SLE were more likely to present latent PolyA for RA, AITD, type 1 diabetes mellitus, and antiphospholipid syndrome *(18)*. Factors such as age and gender were associated with the development of this type of PolyA *(18)*. As previously revised *(19)*, PolyA “may represents the effect of a single genotype and similar environmental factors on diverse phenotypes, and it is associated with female gender, familial autoimmunity, Amerindian ancestry, and cigarette smoking”. All above mentioned data indicate that PolyA is a complex trait in which multiple etiological factors converge.

Herein, we present a yearlong study in three tertiary centers in Colombia, which allowed to classify PolyA, and develop a biomarker model based on autoantibodies positivity. Moreover, we aimed to estimate the frequency of latent and overt PolyA in RA, SLE, SS, AITD, and SSc, along with immunological characteristics.

## RESULTS

### Clinical manifestations are shared among autoimmune diseases due to polyautoimmunity

The general characteristics of patients are shown in Table 1 *(20)*. Most patients included were women. Patients with SLE were younger and exhibited an earlier age at onset. Familial autoimmunity was equally distributed among diseases. Patients with SLE disclosed the highest rates of management with corticosteroids, disease-modifying antirheumatic drugs (DMARDs), and antimalarials, whereas patients with RA reported the highest rates of management with biologics.

**Table 1.** General characteristics of patients with autoimmune diseases.

| Variables (%) | RA (n: 146) | SLE (n: 45) | SS (n: 29) | AITD (n: 17) | SSc (n: 17) | P value <sup>a</sup> |
| --- | --- | --- | --- | --- | --- | --- |
| **Sociodemographic** |  |  |  |  |  |  |
| Age (Median – IQR) | 55 (44.3 – 62) | 39 (32 – 49) | 55 (45 – 61) | 57 (42 – 60) | 54 (47 – 62) | < 0.0001 |
| Sex |  |  |  |  |  | 0.1424 |
| Male | 21 (14.4%) | 7 (15.6%) | 0 (0.0%) | 2 (11.8%) | 1 (5.9%) |  |
| Female | 125 (85.6%) | 38 (84.4%) | 29 (100.0%) | 15 (88.2%) | 16 (94.1%) |  |
| Age at onset (Median – IQR) | 41 (34 – 51) | 29 (24 – 44) | 46 (35.5 – 57) | 35 (27 - 40) | 46 (42 – 49) | < 0.0001 |
| Familial autoimmunity <sup>b</sup> | 37 (25.3%) | 10 (22.2%) | 8/28 (28.6%) | 4 (23.5%) | 2 (11.8%) | 0.7846 |
| Familial autoimmune disease <sup>b</sup> | 23 (15.8%) | 5 (11.1%) | 2/28 (7.1%) | 1 (5.9%) | 1 (5.9%) | 0.6422 |
| Overt polyautoimmunity | 37 (25.3%) | 18 (40.0%) | 5 (17.2%) | 14 (82.4%) | 3 (17.6%) | 0.0005 |
| **Severity of symptoms (Median – IQR) <sup>c</sup>** | 2.9 (2.28 – 4) | 8.5 (4 – 17) | 14 (10 – 19.3) | - | 53.5 (45 – 74.8) | - |
| **Treatment** |  |  |  |  |  |  |
| Corticosteroids | 65 (44.5%) | 30 (66.7%) | 4 (13.8%) | 3 (17.6%) | 3 (17.6%) | 0.0005 |
| DMARDs | 100 (68.5%) | 36 (80.0%) | 3 (10.3%) | 8 (47.1%) | 6 (35.3%) | 0.0005 |
| Antimalarials | 32 (21.9%) | 40 (88.9%) | 5 (17.2%) | 8 (47.1%) | 1 (5.9%) | 0.0005 |
| Biologics | 24 (16.4%) | 3 (6.7%) | 1 (3.4%) | 0 (0.0%) | 0 (0.0%) | 0.0370 |
| **Clinical manifestations** |  |  |  |  |  |  |
| Photophobia | 13/142 (9.2%) | 9/43 (20.9%) | 6/16 (37.5%) | 3 (17.6%) | 0/9 (0.0%) | 0.0110 |
| Malar rash | 2/142 (1.4%) | 9/43 (20.9%) | 0/16 (0.0%) | 0 (0.0%) | 0/9 (0.0%) | 0.0010* |
| Raynaud | 10/142 (7.0%) | 15/43 (34.9%) | 3/16 (18.8%) | 2 (11.8%) | 8/9 (88.9%) | 0.0005* |
| Arthritis | 124/142 (87.3%) | 15/43 (34.9%) | 2/16 (12.5%) | 8 (47.1%) | 4/9 (44.4%) | 0.0005* |
| Arthralgias | 137/142 (96.5%) | 30/43 (69.8%) | 13/16 (81.2%) | 15 (88.2%) | 5/9 (55.6%) | 0.0005* |
| Xerophthalmia | 72/142 (50.7%) | 20/43 (46.5%) | 15/16 (93.8%) | 13 (76.5%) | 4/9 (44.4%) | 0.0045 |
| Xerostomia | 66/142 (46.5%) | 19/43 (44.2%) | 14/16 (87.5%) | 8 (47.1%) | 4/9 (44.4%) | 0.0230 |
| Chronic kidney disease | 2/142 (1.4%) | 7/43 (16.3%) | 0/16 (0.0%) | 0 (0.0%) | 0/9 (0.0%) | 0.0045 |
| Oral ulcers | 4/142 (2.8%) | 7/43 (16.3%) | 2/16 (12.5%) | 1 (5.9%) | 2/9 (22.2%) | 0.0030 |
| Periodontal disease | 6/142 (4.2%) | 1/43 (2.3%) | 4/16 (25.0%) | 1 (5.9%) | 0/9 (0.0%) | 0.0340 |
| Skin ulcers | 0/142 (0.0%) | 1/43 (2.3%) | 0/16 (0.0%) | 0 (0.0%) | 2/9 (22.2%) | 0.0020 |
| Anemia | 16/142 (11.3%) | 15/43 (34.9%) | 5/16 (31.2%) | 2 (11.8%) | 1/9 (11.1%) | 0.0045 |
| Telangiectasias | 5/142 (3.5%) | 0/43 (0.0%) | 1/16 (6.2%) | 1 (5.9%) | 8/9 (88.9%) | 0.0005* |
| Pleural effusion | 1/142 (0.7%) | 7/43 (16.3%) | 1/16 (6.2%) | 0 (0.0%) | 0/9 (0.0%) | 0.0015 |
| Pulmonary embolism | 0/142 (0.0%) | 2/43 (4.7%) | 1/16 (6.2%) | 0 (0.0%) | 0/9 (0.0%) | 0.0500 |
| Pericarditis | 0/142 (0.0%) | 1/43 (2.3%) | 0/16 (0.0%) | 0 (0.0%) | 0/9 (0.0%) | 0.3608 |
| Seizures | 1/142 (0.7%) | 0/43 (0.0%) | 0/16 (0.0%) | 1 (5.9%) | 0/9 (0.0%) | 0.3843 |
| Psychosis | 0/142 (0.0%) | 0/43 (0.0%) | 0/16 (0.0%) | 0 (0.0%) | 0/9 (0.0%) | - |
| Vasculitis | 0/142 (0.0%) | 0/43 (0.0%) | 0/16 (0.0%) | 0 (0.0%) | 0/9 (0.0%) | - |
| CNS compromise | 0/142 (0.0%) | 3/43 (7.0%) | 1/16 (6.2%) | 0 (0.0%) | 0/9 (0.0%) | 0.0210 |
| PNS compromise | 0/142 (0.0%) | 1/43 (2.3%) | 0/16 (0.0%) | 0 (0.0%) | 0/9 (0.0%) | 0.3663 |
| Myalgia | 26/142 (18.3%) | 9/43 (20.9%) | 3/16 (18.8%) | 1 (5.9%) | 1/9 (11.1%) | 0.7571 |
| Calcinosis | 1/142 (0.7%) | 0/43 (0.0%) | 0/16 (0.0%) | 0 (0.0%) | 0/9 (0.0%) | 1.0000 |
| Urticaria | 2/142 (1.4%) | 1/43 (2.3%) | 2/16 (12.5%) | 1 (5.9%) | 0/9 (0.0%) | 0.0865 |
| Alopecia | 12/142 (8.5%) | 11/43 (25.6%) | 1/16 (6.2%) | 4 (23.5%) | 1/9 (11.1%) | 0.0255 |
| Dysphagia | 4/142 (2.8%) | 1/43 (2.3%) | 3/16 (18.8%) | 3 (17.6%) | 4/9 (44.4%) | 0.0005* |
| Gastritis | 20/142 (14.1%) | 5/43 (11.6%) | 8/16 (50.0%) | 3 (17.6%) | 1/9 (11.1%) | 0.0105 |
| Weight loss | 4/142 (2.8%) | 3/43 (7.0%) | 3/16 (18.8%) | 0 (0.0%) | 1/9 (11.1%) | 0.0415 |
| Infertility | 1/142 (0.7%) | 1/43 (2.3%) | 0/16 (0.0%) | 1 (5.9%) | 0/9 (0.0%) | 0.2974 |
| Periorbital edema | 1/142 (0.7%) | 1/42 (2.4%) | 1/16 (6.2%) | 1 (5.9%) | 0/9 (0.0%) | 0.1479 |
| Sclerodactyly | 1/142 (0.7%) | 0/43 (0.0%) | 0/16 (0.0%) | 1 (5.9%) | 8/9 (88.9%) | 0.0005* |
| Miscarriage | 19/123 (15.4%) | 8/36 (22.2%) | 3/16 (18.8%) | 4/15 (26.7%) | 1/8 (12.5%) | 0.6822 |
| Preeclampsia | 8/123 (6.5%) | 2/36 (5.6%) | 0/16 (0.0%) | 2/15 (13.3%) | 0/8 (0.0%) | 0.6682 |
| Preterm delivery | 9/123 (7.3%) | 3/36 (8.3%) | 1/16 (6.2%) | 0/15 (0.0%) | 0/8 (0.0%) | 0.9245 |
| Vascular thrombosis | 2/142 (1.4%) | 5/43 (11.6%) | 1/16 (6.2%) | 1 (5.9%) | 1/9 (11.1%) | 0.0140 |
| Episcleritis | 1/142 (0.7%) | 0/43 (0.0%) | 0/15 (0.0%) | 0 (0.0%) | 0/9 (0.0%) | 1.0000 |
| Uveitis | 0/142 (0.0%) | 0/43 (0.0%) | 0/16 (0.0%) | 0 (0.0%) | 0/9 (0.0%) | - |
| Skin nodules | 4/142 (2.8%) | 1/43 (2.3%) | 0/16 (0.0%) | 1 (5.9%) | 0/9 (0.0%) | 0.8136 |
| Pulmonary nodules | 0/142 (0.0%) | 0/43 (0.0%) | 0/16 (0.0%) | 0 (0.0%) | 1/9 (11.1%) | 0.0455 |
| Goiter | 4/142 (2.8%) | 0/43 (0.0%) | 0/16 (0.0%) | 4 (23.5%) | 0/9 (0.0%) | 0.0070 |
| Menstrual disorders | 33/123 (26.8%) | 13/36 (36.1%) | 5/16 (31.2%) | 5/16 (31.2%) | 1/8 (12.5%) | 0.7141 |
| Morphea | 0/142 (0.0%) | 0/43 (0.0%) | 0/16 (0.0%) | 0 (0.0%) | 0/9 (0.0%) | - |
| Nail dystrophy | 0/142 (0.0%) | 1/43 (2.3%) | 0/16 (0.0%) | 0 (0.0%) | 0/9 (0.0%) | 0.3688 |
| Microstomia | 0/142 (0.0%) | 0/43 (0.0%) | 0/16 (0.0%) | 0 (0.0%) | 1/9 (11.1%) | 0.0395 |
| Heliotrope rash | 0/122 (0.0%) | 0/33 (0.0%) | 0/16 (0.0%) | 0/16 (0.0%) | 0/7 (0.0%) | - |
| Gottron papules | 0/122 (0.0%) | 0/32 (0.0%) | 0/16 (0.0%) | 0/16 (0.0%) | 0/6 (0.0%) | - |
| Hand edema | 3/121 (2.5%) | 0/32 (0.0%) | 0/16 (0.0%) | 1/16 (6.2%) | 2/6 (33.3%) | 0.0235 |
| **Clinical tests <sup>†</sup>** |  |  |  |  |  |  |
| ESR mm/hr (Median – IQR) | 25 (14 - 36) | 21 (14 - 36.5) | 24 (13.5 - 36.5) | 21 (11 - 37) | 26 (19 - 35) | 0.9436 |
| CRP mg/dL (Median – IQR) | 0.6 (0.3 - 1.5) | 0.5 (0.3 - 1.6) | 0.5 (0.3 - 1.2) | 0.4 (0.3 - 0.7) | 0.5 (0.4 - 0.8) | 0.7438 |
| Fibrinogen mg/dL (Median – IQR) | 352 (284 – 479) | 382 (283.5 - 467.5) | 314 (261.5 - 442.3) | 286 (257 - 338) | 363.5 (277 - 476.3) | 0.2501 |
| TSH μIU/mL (Median – IQR) | 2.2 (1.4 - 4.0) | 3.0 (1.6 - 3.43) | 2.3 (1.4 - 4.0) | 3.3 (1.8 - 4.3) | 4.6 (4.5 - 5.0) | 0.3143 |
| Leukocytes (Median – IQR) | 7,050 (5,773 – 8,025) | 4,470 (3,508 - 6,475) | 6,100 (4,450 – 6,750) | 5,250 (4,078 – 6,425) | 6440 (5,720 – 6,520) | < 0.0001 |
| Lymphocytes (Median – IQR) | 2,000 (1,600 – 2,500) | 1,200 (900 – 1,570) | 1,700 (1,400 – 2,250) | 1,380 (1,130 – 1,786.3) | 1600 (1,565 – 1,600) | < 0.0001 |
| Hemoglobin g/dL (Median – IQR) | 13.7 (12.7 - 14.5) | 12.50 (11.2 - 13.3) | 13.3 (12.0 - 14.8) | 13.5 (12.2 - 14.5) | 13.7 (11.4 - 13.7) | 0.0219 |

<sup>a</sup> Quantitative variables were analyzed by Kruskal-Wallis test, whereas qualitative variables were analyzed by Fisher's exact test.
<sup>b</sup> Familial autoimmunity corresponds to the presence of different autoimmune diseases in a nuclear family. On the other hand, familial autoimmune disease is defined as the presence of one specific autoimmune disease in various members of a nuclear family (i.e., coaggregation).<sup>36</sup>
<sup>c</sup> Severity of symptoms was evaluated as follows: DAS-28 in RA, SLAQ in SLE, ESSPRI in SS and SSPro in SSc.
<sup>†</sup> For ESR and CRP: 143 patients for RA; 17 AITD; 9 SSc; 43 SLE; 16 SS. For fibrinogen: 141 RA; 17 AITD; 8 SSc; 43 SLE; 16 SS. For TSH: 79 for RA; 9 for AITD; 3 for SSc; 18 for SLE; 12 for SS. For leukocytes, lymphocytes, and hemoglobin: 80 for RA; 14 for AITD; 3 for SSc; 42 for SLE; 11 for SS.
\* Statistically significant after Bonferroni correction. P value threshold for clinical manifestations: 0.05/41= 0.0012.
RA: Rheumatoid arthritis; SLE: Systemic lupus erythematosus; SS: Sjögren's syndrome; AITD: Autoimmune thyroid disease; SSc: Systemic sclerosis; DAS-28: Disease activity score 28; SLAQ: Systemic lupus activity questionnaire; ESSPRI: EULAR Sjogren's syndrome patient reported index; SSPro: Scleroderma skin patient reported outcome; DMARDs: disease-modifying antirheumatic drugs; ESR: Erythrocyte sedimentation rate; CRP: C reactive protein; TSH: Thyroid stimulating hormone; CNS: Central nervous system, PNS: Peripheral nervous system; IQR: interquartile range; NA: Not applicable/Available.

Patients with RA presented low activity of disease, according to DAS-28. On the contrary, most patients with SLE showed moderate clinical reported activity (i.e., SLAQ score ≥ 3). Raynaud, telangiectasias, dysphagia, and sclerodactyly were most common in SSc. Arthritis and arthralgias were most frequently presented in RA, whereas malar rash was distinctive of SLE (Table 1). Interestingly, some clinical manifestations were equally distributed across diseases, probably due to PolyA. Clinical inflammatory biomarkers (i.e., ESR, CRP and fibrinogen) and thyroid function did not differ among diseases. However, patients with SLE disclosed lower levels of total leukocytes, lymphocytes, and hemoglobin (Table 1).

### Patients with autoimmune diseases share expression of IgG autoantibodies

Initially, we evaluated the expression of autoantibodies compared with 38 healthy volunteers. After quality control filtering (i.e., SNR > 3), 111 IgG and 97 IgM autoantibodies were included in the final analysis. It was found that 25 IgG autoantibodies in SLE, 7 in SS, 2 in AITD, and 12 in SSc were over-expressed (i.e., Log2 fold change ≥ 1, and p-value FDR <0.05) (Supplementary Material). There were no over-expressed nor under-expressed IgG autoantibodies in patients with RA (Fig. 1A) and this finding was related to the lack of rheumatoid factor (RF) and citrullinated antigens included in the microarray. Patients with different index ADs shared several IgG autoantibodies (i.e., PolyA) (Fig. 1B).

**Fig. 1.**
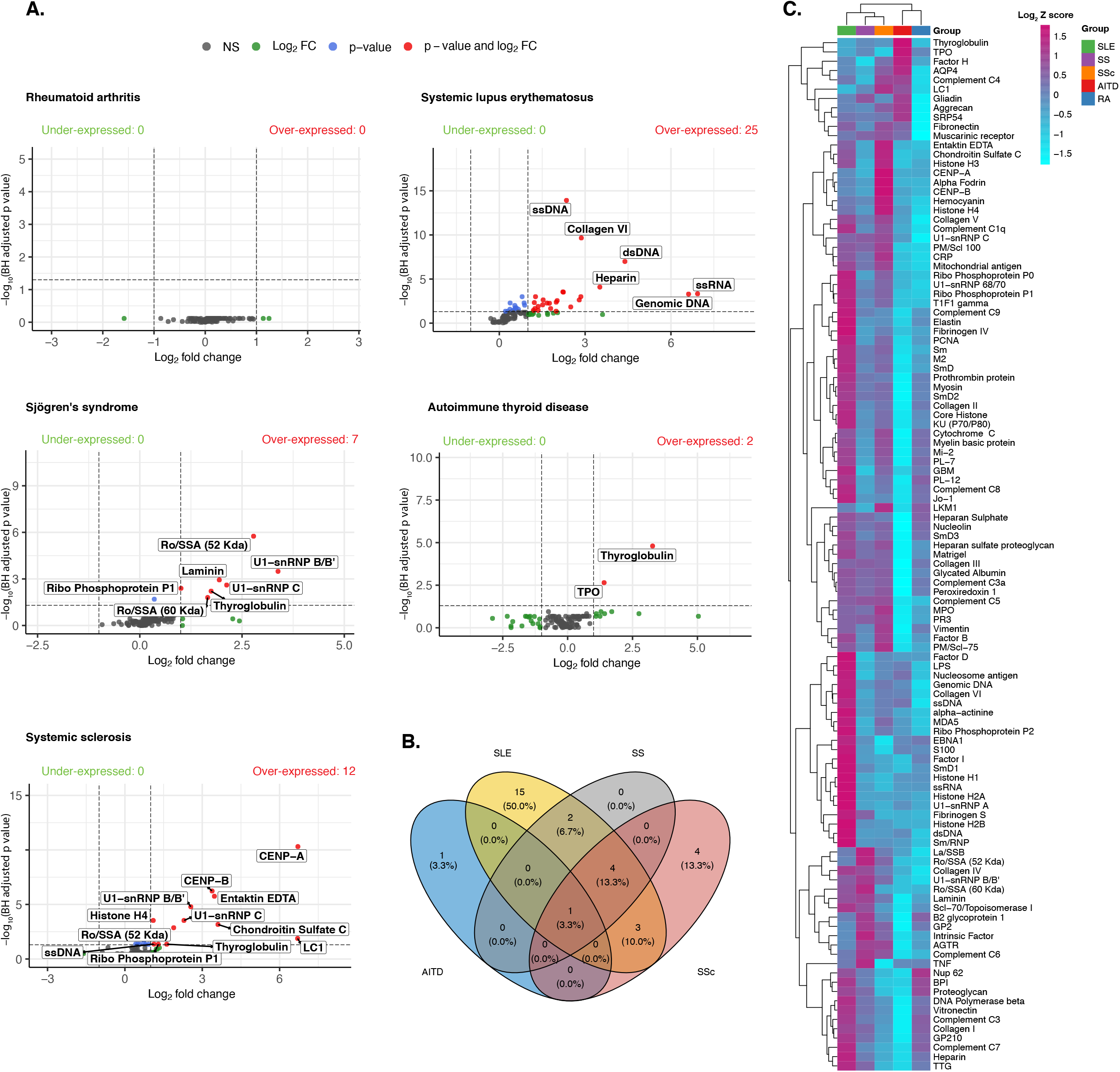
IgG microarray analysis. **(A)** Volcano plots for IgG autoantibodies in each condition. Red dots represent those autoantibodies with Log2 fold change ≥ 1, and p-value FDR <0.05. Analysis included 146 patients with RA, 45 with SLE, 29 with SS, 17 with AITD, and 17 with SSc. **(B)** Venn diagram for overexpressed IgG autoantibodies shared among diseases. **(C)** Heatmap for 111 IgG autoantibodies. The color of the heatmap varies from blue, which indicates under-expression, to purple, which indicates over-expression. Clustering was performed using the ward agglomeration method. NS: Not significant; FC: Fold change, FDR: False discovery rate; BH: Benjamín-Hochberg; RA: Rheumatoid arthritis; SLE: Systemic lupus erythematosus; SS: Sjögren’s syndrome; AITD: Autoimmune thyroid disease; SSc: Systemic sclerosis.

Patients with SLE showed over-expression of IgG autoantibodies against nuclear, thyroid, complement and collagen-associated antigens (Fig. 1C). This was similar for SS, in which autoantibodies for ribonuclear proteins and thyroid antigens were over-expressed. Concerning AITD, only IgG autoantibodies against thyroid antigens were significantly over-expressed. Patients with SSc showed over-expression of autoantibodies against nuclear, cytoplasmatic and thyroid antigens (Fig. 1C).

Few IgM autoantibodies were over-expressed in ADs (Supplementary Appendix) (Fig. 2A) and sharing of IgM autoantibodies was less likely (Fig. 2B). IgM autoantibodies against nuclear antigens in AITD and SLE were over-expressed. Interestingly, patients with SSc showed under-expression of autoantibodies for nuclear and myelinic antigens, but over-expression of liver associated autoantibodies (i.e., LC1) (Fig. 2C). There were not under-expressed nor over-expressed IgM autoantibodies in patients with RA and SS.

**Fig. 2.**
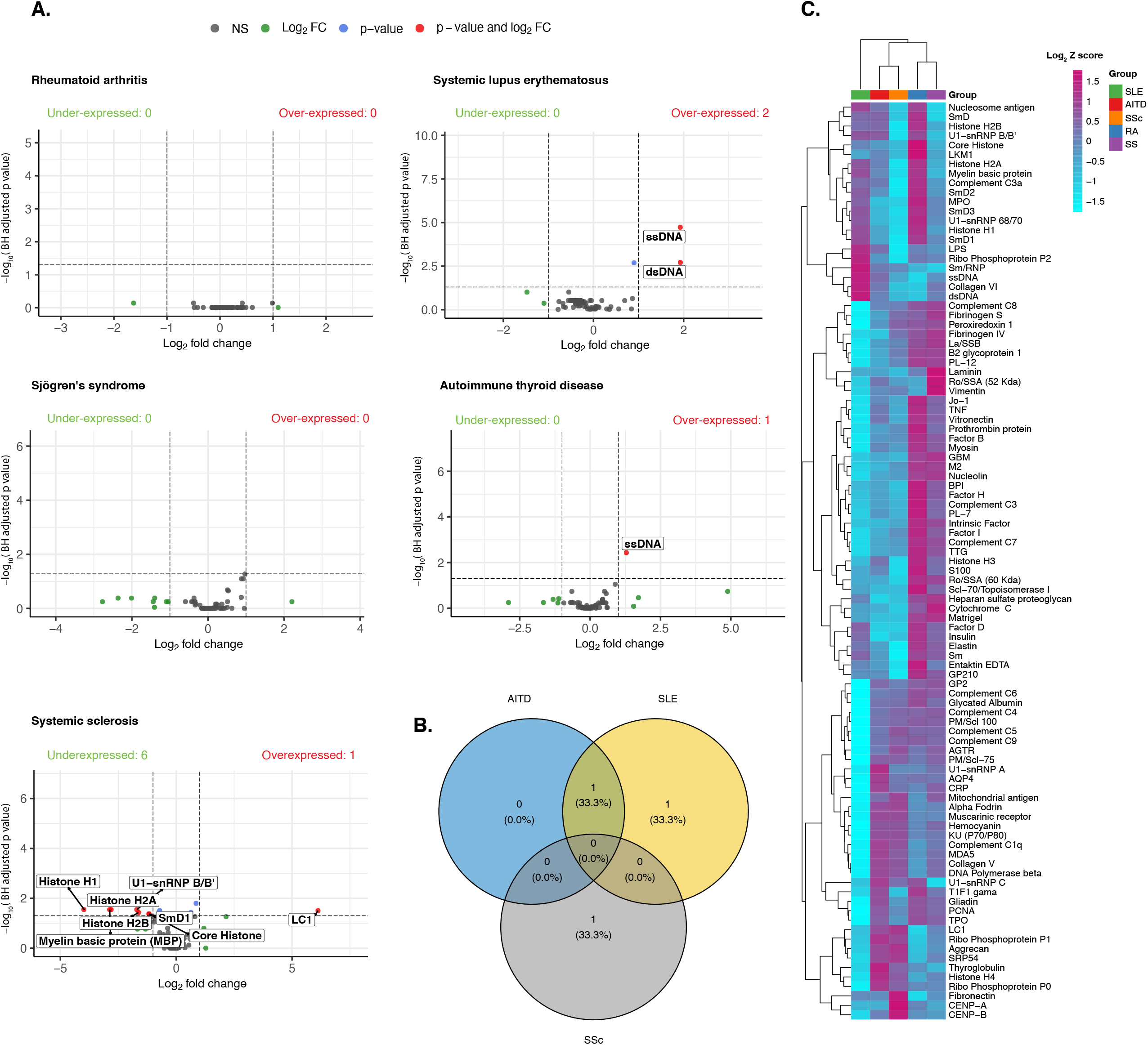
IgM microarray analysis. **(A)** Volcano plots for IgM autoantibodies in each condition. Red dots represent those autoantibodies with Log2 fold change ≥ 1, and p-value FDR <0.05. Analysis included 146 patients with RA, 45 with SLE, 29 with SS, 17 with AITD, and 17 with SSc. **(B)** Venn diagram for overexpressed IgM autoantibodies shared among diseases. **(C)** Heatmap for 97 IgM autoantibodies. The color of the heatmap varies from blue, which indicates under-expression, to purple, which indicates over-expression. Clustering was performed using the ward agglomeration method. NS: Not significant; FC: Fold change, FDR: False discovery rate; BH: Benjamín-Hochberg; RA: Rheumatoid arthritis; SLE: Systemic lupus erythematosus; SS: Sjögren’s syndrome; AITD: Autoimmune thyroid disease; SSc: Systemic sclerosis.

### IgG autoantibodies yield high accuracy for classification of autoimmune diseases

Next, we focused on the evaluation of over-expressed IgG autoantibodies (i.e., Log2 fold change ≥ 1, and p-value FDR <0.05) as hallmarks for the identification of index AD. To avoid the bias of patients with overt PolyA, this analysis only those patients without such condition. Multivariate logistic regression yielded that IgG autoantibodies against nuclear antigens disclosed the best performance in SLE (Table 2). In AITD, IgG against Tg was the only associated autoantibody. Tg was associated with classification of SS and SSc. In addition, autoantibodies against ribonucleoproteins and centromere were also associated with classification of SS and SSc, respectively (Fig. 3A).

**Table 2.**
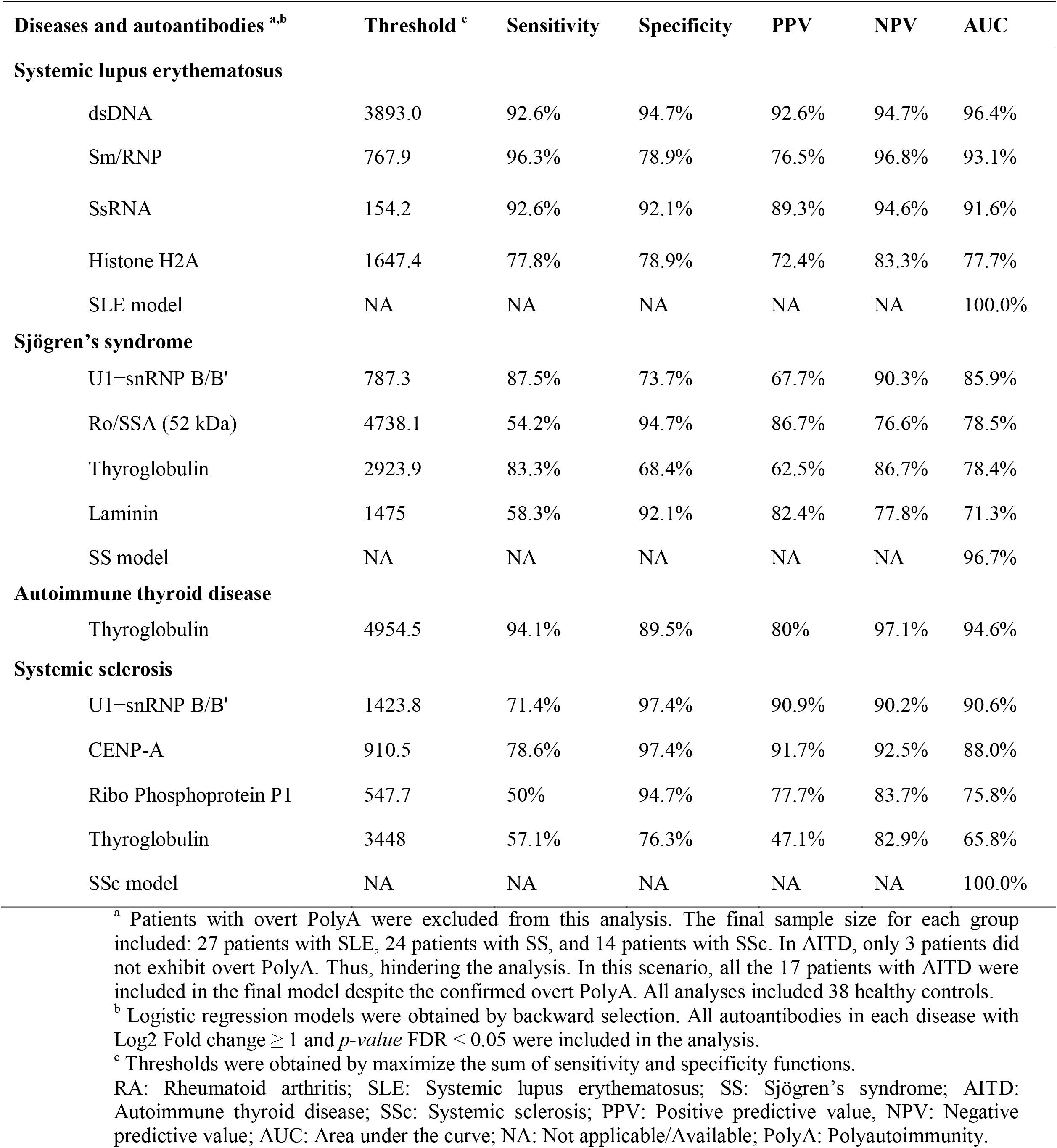
Diagnostic accuracy of autoantibodies selected by multivariate analysis.

**Fig. 3.**
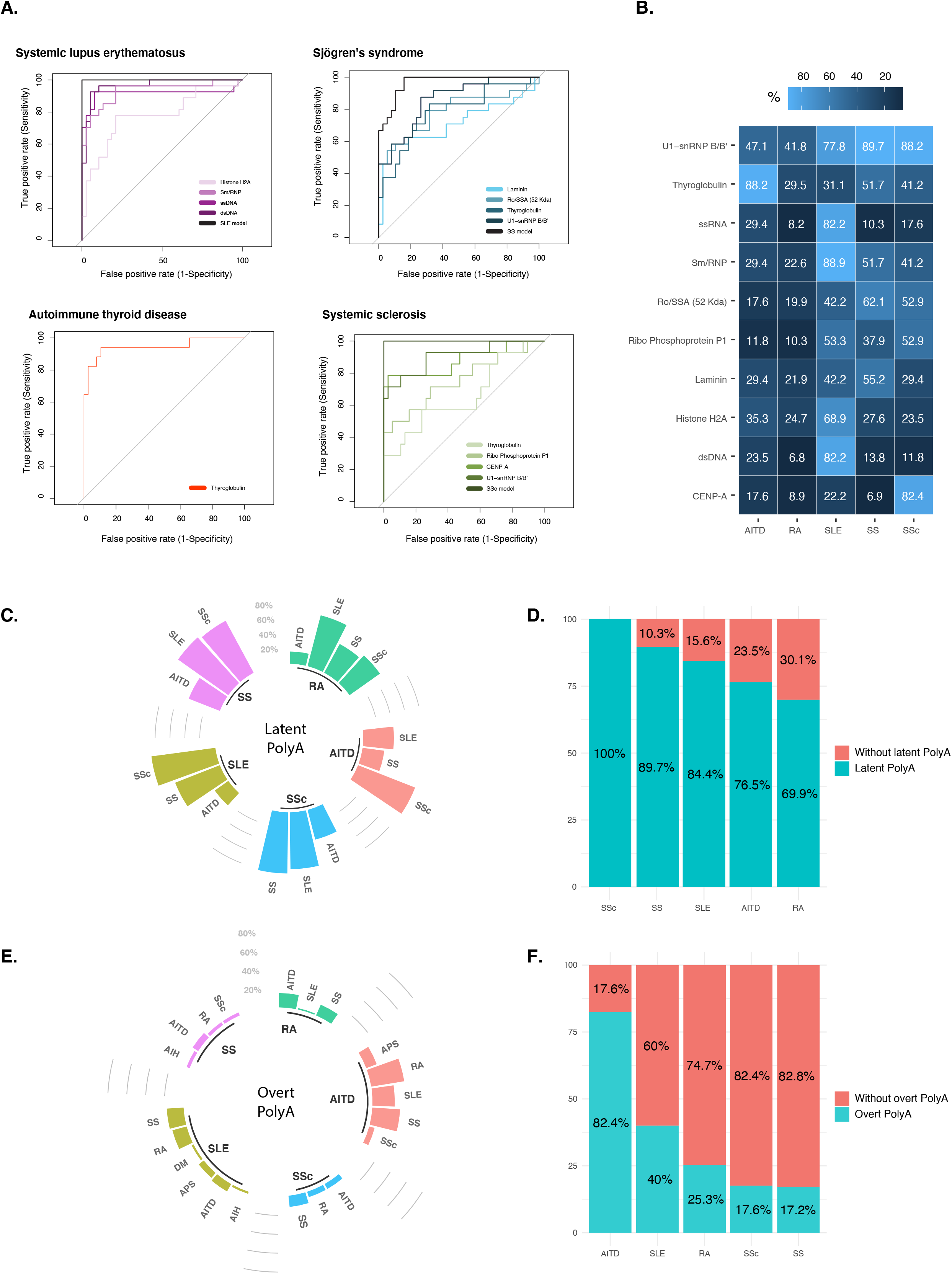
Prevalence of latent and overt PolyA. **(A)** ROC curves for selected IgG autoantibodies from multivariate logistic regression analysis. **(B)** Heat map for positivity of selected IgG autoantibodies. **(C)** Circular bar plot for sources of latent PolyA. **(D)** Bar plot for overall prevalence of latent PolyA. **(E)** Circular bar plot for sources of overt PolyA. **(F)** Bar plot for overall prevalence of overt PolyA. ROC: Receiver operating characteristic; PolyA: Polyautoimmunity; RA: Rheumatoid arthritis; SLE: Systemic lupus erythematosus; SS: Sjögren’s syndrome; AITD: Autoimmune thyroid disease; SSc: Systemic sclerosis; AIH: Autoimmune hepatitis; APS: Antiphospholipid syndrome; DM: Type 1 diabetes mellitus.

### Latent polyautoimmunity surpasses overt polyautoimmunity in autoimmune diseases

Next, given the thresholds of the IgG autoantibodies obtained (Table 2), we evaluated their positivity in all included diseases to estimate the frequency of latent PolyA. Since Tg and U1−snRNP B/B’ IgG autoantibodies were shared in several conditions, only those estimated thresholds for AITD and SS were used, respectively.

Positivity for included autoantibodies in all patients is shown in Figure 3B. Although IgG autoantibodies obtained by multivariate analysis exhibited high frequency in specific index conditions, patients showed positivity for other autoantibodies (i.e., PolyA). The overall frequency of these autoantibodies was low in RA, whereas SLE, SS, and SSc showed higher positivity rates (Fig. 3B).

Then, we looked for those autoantibodies that were not associated with overt ADs to estimate the occurrence of latent PolyA in each condition (Fig. 3C). Latent SLE was the most common in patients with RA and SS. On the other hand, latency for SSc was the most common in patients with AITD and SLE, whereas latency for AITD was the most common in SSc. This analysis yielded that more than 70% of patients presented at least 1 type of latency for SLE, SS, AITD, or SS (Fig. 3D).

Based on classification criteria, the frequency of overt PolyA was estimated. AITD was the most common cause of overt PolyA in SS and RA (Fig. 3E). Conversely, RA was the most common cause of overt PolyA in AITD. In patients with SLE and SSc, overt PolyA was predominantly defined by SS. In contrast to latent PolyA, overt PolyA was less frequent, and AITD presented the highest rates (Fig. 3F).

### Polyautoimmunity influences clinical and immunological phenotypes in autoimmune diseases

After estimation of the occurrence of overt and latent PolyA, we developed a classification of ADs based on autoantibodies. Three main clusters were obtained (Fig. 4A-B). Clusters 2 and 3 exhibited the highest frequency of latent (Fisher’s exact test, P= 0.0005) (Fig. 4C), and overt PolyA (Fisher’s exact test, P= 0.0025) (Fig. 4E). SSc and SS were the most common cause of latent and overt PolyA in both clusters, respectively (Fig. 4D-F).

**Fig. 4.**
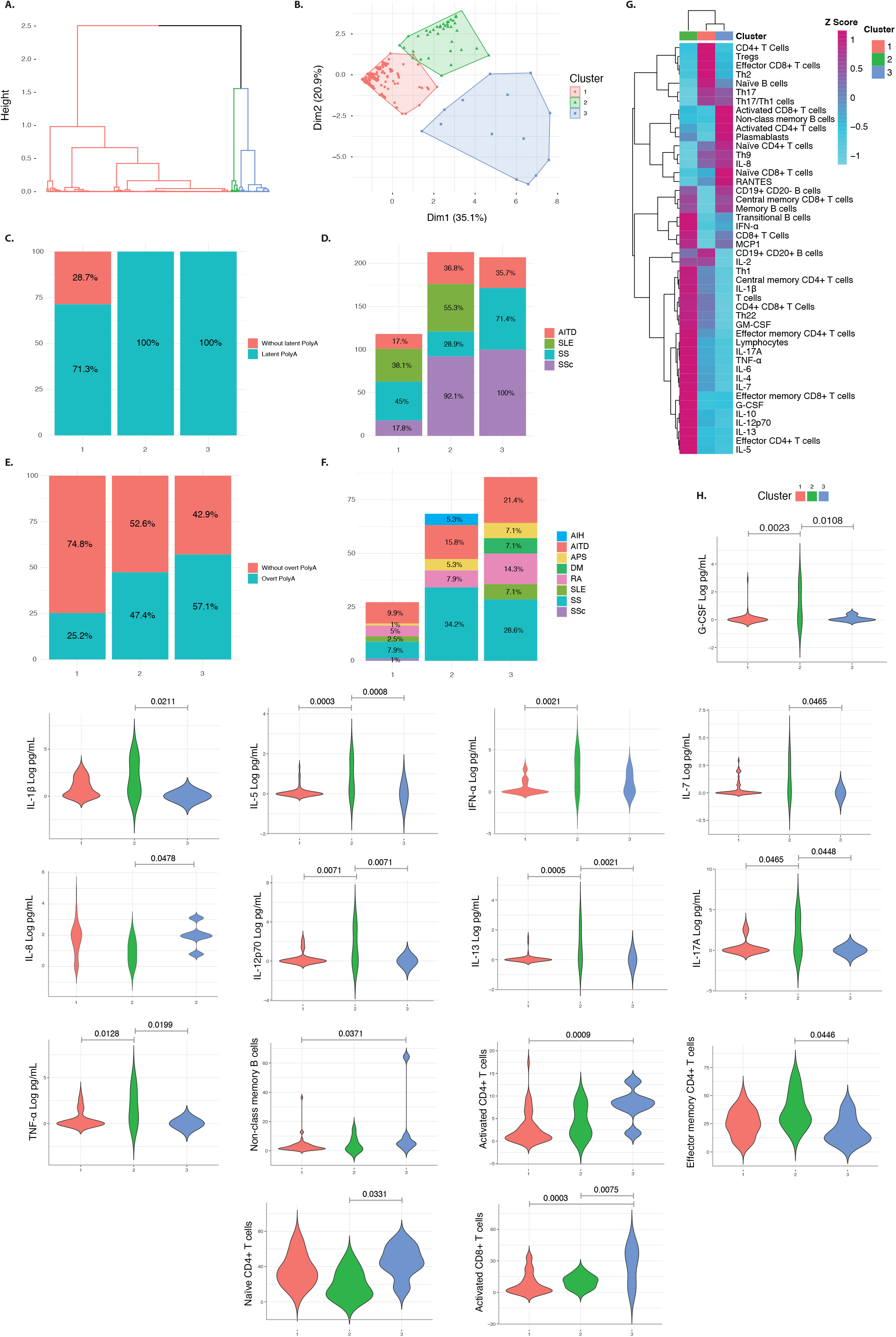
PolyA-based classification of ADs. **(A)** Cluster dendrogram for classification of ADs based on selected autoantibodies. **(B)** Factor map for obtained clusters. **(C)** Bar plot for overall prevalence of latent PolyA by cluster. **(D)** Bar plot for sources of latent PolyA by cluster. **(E)** Bar plot for overall prevalence of overt PolyA by cluster. **(F)** Bar plot for sources of overt PolyA by cluster. **(G)** Heatmap for cytokines and lymphocytes immunophenotype. Analysis included 67 patients. The color of the heatmap varies from blue, which indicates under-expression, to purple, which indicates over-expression. Clustering was performed using the ward agglomeration method. The log-transformed cytokine concentration was used to construct the heatmap. **(H)** Representative violin plots of cytokines and lymphocytes phenotypes. Statistical analysis was performed using linear models that were adjusted for age, and sex. G-CSF: Granulocyte colony-stimulating factor; IL: Interleukin; TNF-α: Tumor necrosis factor-alpha; IFN-α: Interferon-alpha; RANTES: Regulated on activation, normal T cell expressed and secreted; MCP-1: Monocyte chemotactic protein-1; PolyA: Polyautoimmunity; RA: Rheumatoid arthritis; SLE: Systemic lupus erythematosus; SS: Sjögren’s syndrome; AITD: Autoimmune thyroid disease; SSc: Systemic sclerosis; AIH: Autoimmune hepatitis; APS: Antiphospholipid syndrome; DM: Type 1 diabetes mellitus.

Interestingly, cluster 3 was most likely to receive treatment with corticosteroids, DMARDs, and immunosuppressors (Fisher’s exact test, P<0.0105), and presented higher frequency of malar rash, Raynaud, oral ulcers, and central nervous system compromise (Fisher’s exact test, P<0.0500). On the other hand, cluster 2 was characterized by xeropthalmia, xerostomia, periodontal disease, and weight loss (Fisher’s exact test, P<0.0500).

Cytokine and lymphocyte profiles were different among clusters (Fig. 4G). Cluster 2 exhibited a dysregulated immunological profile given by high levels of G-CSF, IL-5, INF-α, IL-7, IL-12p70, IL-13, IL-17A, TNF-α, and effector memory CD4+ T cells, whereas naïve CD4+ T cells were decreased (Fig. 4H). On the other hand, Cluster 3, showed high levels of IL-8, activated CD4+ and CD8+ T cells (Fig.4H).

## DISCUSSION

In this proof-of-concept study, it is confirmed that PolyA allows a new taxonomy of ADs. Clusters of PolyA were well differentiated and characterized by a unique immune signature (i.e., cytokine response and cellular subphenotypes) (Fig. 4). Moreover, latent PolyA was more frequent than overt PolyA *(2, 3, 5)*.

Several autoantibodies share different specificities across ADs. For example, anti-SSA/Ro and anti-SSB/La are considered the two most classic autoantibodies in SS *(6)*, and nearly 63% of patients show positivity to anti-SSA/Ro *(21)*. However, this autoantibody was also associated with the development of SLE *(22)*. Anti-SSA/Ro in the presence of anti-SSB/La tends to identify patients with SS. It was found that 29 out of 35 patients with both anti-Ro/SSA and anti-La/SSB had SS, whereas of 53 with only anti-Ro/SSA, 23 had SS, 25 had SLE, and 13 had another disease *(23)*. This suggests that the combination of some autoantibodies in the diagnostic approach of ADs may improve the sensitivity and specificity of these tests *(3)*.

The microarray analysis allowed the identification of mixtures of autoantibodies that helped to develop high accuracy models for classification of ADs. Thus, confirming the usefulness of antibody combination in the diagnosis of disease. Tg and U1−snRNP B/B’ autoantibodies were shared as diagnostic biomarkers for AITD, SS and SSc. This may suggest that some autoantibodies yielded similar specificities across diseases, or a phenomenon of latent PolyA, in which these patients may develop overt PolyA in the future. Usefulness of the models obtained deserves further attention and confirmation in larger studies.

The frequency of latent and overt PolyA matches with prior studies in which different causes of PolyA were described *(1–3)*. In this study, we found combinations of diverse ADs conforming the spectrum of PolyA. Latent PolyA was most common than overt PolyA. This may suggest that most patients with an index condition present autoantibody positivity for other ADs, and thus inferring that primary or secondary labels of ADs are inaccurate. In this line, patients with any ADs should be tested for other types of PolyA. This may have implications for follow-up and treatment (i.e., primary prevention). As mentioned, positivity for autoantibodies predate the appearance of overt ADs *(6)*, and in SSc, it has been suggested that PolyA may influence deleterious outcomes such as pulmonary fibrosis and mortality *(24, 25)*. As a corollary PolyA should be considered in all studies dealing with ADs, including epidemiological, genetic, and clinical trials.

The possible shortcomings of our study must be acknowledged. The main objective of this cross-sectional study was to estimate the frequency of latent and overt PolyA in outpatient clinics. In this line, this study reflects the frequency of ADs in our settings, being RA the most frequent, and supporting the unequal final sample size per group. In addition, patients with RA did not show over-expressed autoantibodies. The lack of RF and citrullinated antigens included in the autoantigen array chip could be associated with these results.

Further studies implementing these antigens are warranted. Frequency latencies were estimated only for those ADs included. Thresholds and specificities for other ADs were not estimated (e.g., gastrointestinal or endocrinological ADs). It is necessary to test this microarray in other ADs, to assess thresholds of positivity, and to improve the clinical efficiency of this technique. It is likely that tests for positivity for other ADs would have expanded PolyA, and latent PolyA would have reached higher rates.

## MATERIALS AND METHODS

### Study design and patients’ recruitment

A cross-sectional study was conducted from December 1st, 2018, to November 30th, 2019, in three tertiary specialized rheumatology centers; two were in Bogota, Colombia, including the Dermatology and Rheumatology Foundation (FUNINDERMA), and the Center for Autoimmune Diseases Research (CREA). The remaining center was the “Centro de Referencia en Osteoporosis, Reumatología & Dermatología”, in Cali, Colombia.

Two-hundred and eighty-one consecutive patients attending the outpatient clinic were assessed. Only those patients with the following index conditions were considered: RA, SLE, SS, AITD, and SSc. The patients fulfilled either the 1987 American College of Rheumatology (ACR) classification criteria for RA *(26)*, the 1997 ACR criteria for SLE *(27)*, the 2013 ACR/European League Against Rheumatism (EULAR) classification criteria for SSc *(28)*, or the revised American-European Consensus Group for SS *(29)*. For AITD, patients with autoimmune hypothyroidism (AH) were classified as follows: 1) confirmed AH (i.e., thyroid dysfunction, thyroid stimulating hormone (TSH) >4.1 μIU/mL or levothyroxine treatment, and the presence of anti-thyroperoxidase (TPO) or anti-thyroglobulin (Tg) antibodies), 2) euthyroid patients with positive anti-TPO or anti-Tg antibodies, 3) non-autoimmune hypothyroidism (thyroid dysfunction and absence of anti-TPO or anti-Tg antibodies) *(30)*.

Then, foreign patients (n: 2), patients with prior history of neoplasia (n: 4), and unfulfillment of classification criteria (n: 21) were excluded. A final sample size of 254 patients was included in the analyses as follows: RA (n: 146), SLE (n: 45), SS (n: 29), AITD (n: 17) and SSc (n: 17). In addition, a group of 38 healthy volunteers (i.e., subjects without overt autoimmunity nor familial autoimmunity) were included as a control group.

This study was done in compliance with the Act 008430/1993 of the Ministry of Health of the Republic of Colombia, which classified it as minimal-risk research. All the patients were asked for their consent and were informed about the Colombian data protection law (1581 of 2012). The institutional review board of the Universidad del Rosario approved the study design.

### Patient monitoring and clinical evaluation

The patients’ demographic and cumulative clinical data were simultaneously obtained by standardized report form, physical examination, and chart review. Data included age, age at onset of disease, familial autoimmunity and familial autoimmune disease, and treatment on inclusion. All patients were evaluated for rheumatological or associated autoimmune clinical manifestations. These variables are described in Table 1.

In addition, if any patient after inclusion presented positivity for autoantibodies by ELISA and Immunoblot not related to the index condition, clinicians evaluated the subject once again, and performed clinical tests to confirm classification criteria for overt PolyA. These included Schirmer and unstimulated saliva flow rate test for SS, and TSH for AITD. Patients with positivity for anti-phospholipid antibodies were tested within 12 weeks to confirm the classification criteria.

Severity of symptoms was assessed by either disease activity score 28 (DAS-28) for RA *(31)*, systemic lupus activity questionnaire (SLAQ) for SLE *(32)*, scleroderma skin patient-reported outcome (SSPRO) for SSc *(33, 34)*, or EULAR SS patient reported index (ESSPRI) for SS *(34–36)*. In addition, most of the patients were systematically evaluated for erythrocyte sedimentation rate (ESR), C reactive protein (CRP), fibrinogen, and blood count. All data were collected in an electronic and secure database as described elsewhere *(34)*.

### Microarray autoantibody profiling

Samples were analyzed by an autoantigen array chip containing 128 antigens and controls at the Microarray and Immune Phenotyping Core Facility, UT Southwestern Medical Center. Briefly, the autoantigens and control proteins are printed in duplicates onto nitrocellulose film slides. Serum samples were pretreated with DNAse-I and diluted in phosphate buffered saline with Tween (PBST) for autoantibody profiling. The diluted serum samples were incubated with the autoantigen arrays, and autoantibodies binding with antigens on arrays were measured with cy3-conjugated anti-human IgG (Jackson ImmunoResearch Laboratories) and cy5-conjugated anti-human IgM (Jackson ImmunoResearch Laboratories), using a Genepix 4200A scanner (Molecular Device). The resulting images were analyzed with Genepix Pro 7.0 software (Molecular Devices). The median of the signal intensity for each spot was calculated and subtracted from the local background around the spot, and data obtained from duplicated spots were averaged. The background subtracted signal intensity of each antigen was normalized to the average intensity of the human IgG or IgM controls, which were spotted on the array as internal controls. Finally, the normalized fluorescence intensity (NFI) was generated as a quantitative measurement of the binding capacity of each antibody with the corresponding autoantigen. Signal-to-noise ratio (SNR) is another quantitative measurement of the true signal above background noise. SNR values equal to or greater than 3 were considered significantly higher than background, and therefore as true signals. The autoantibody which has the SNR value of less than 3 in more than 90% of the samples was considered negative and excluded from further analysis.

### Cytokine assay and lymphocytes immunophenotype

Serum of patients was collected in fasting state and spite of the treatment status. Concentration of 19 cytokines (IL-2, IL-4, IL-5, IL-6, IL-7, IL-8, IL-9, IL-10, IFN-α, TNF-α, G-CSF, GM-CSF, RANTES, MCP1, IL-12p70, IL-13, IFN-γ) in serum samples from patients were assessed by Cytometric Bead Array (CBA, Becton Dickinson Biosciences, San Diego, CA, USA). The test was done according to the manufacturer’s protocols. Concentration of the cytokines was calculated using the FCAP Array™ Software (BD Bioscience) as reported elsewhere *(5)*. For a detailed analysis of the cell phenotype, peripheral blood mononuclear cells were stained with fluorescent antibodies. A minimum of 100,000 lymphocytes per sample were acquired on a FACSCanto II™ flow cytometer (BD Biosciences™). Twenty-eight cell subsets (Supplementary Appendix,) were analyzed with FlowJo software version 9 (BD Biosciences™) as reported elsewhere *(37)*.

### Statistical analysis

Univariate descriptive statistics were performed. Categorical variables were analyzed using frequencies, and quantitative continuous variables were expressed as the median and interquartile range (IQR). The Kruskal-Wallis, or Fisher’s exact tests were used based on the results. Bonferroni correction was used for multiple testing in clinical manifestations.

Initially, we aimed to evaluate the over-expressed autoantibodies compared with healthy controls. First, data from autoantigen array was standardized by a robust linear model as previously described *(38, 39)*. The p-value was determined by unpaired t-test with a Benjamini and Bonferroni-Hochberg False Discovery Rate post-hoc correction (FDR). For all autoantibodies, the only selected were those that fulfilled: 1) p-value with a Benjamini-Hochberg FDR <0.05, and 2) Log2 fold change ≥ 1.

A logistic regression model was fitted to estimate the effect size of significant autoantibodies on ADs classification (i.e., Log2 fold change ≥ 1, and FDR <0.05). The dependent variable of the logistic model was the natural log of the odds of the index AD. The independent variables of the logistic model were selected through a backward selection procedure as previously described *(40)*.

From the selected autoantibodies, thresholds for positivity were obtained by maximizing the sum of sensitivity and specificity functions comparing it with healthy volunteers. Sensitivity, specificity, positive predictive value (PPV), negative predictive value (NPV), and area under the curve (AUC) were estimated for each threshold (https://github.com/thie1e/cutpointr).

Next, we aimed to develop a new ADs classification based on those autoantibodies selected in the previous step. We used the mixed-cluster methodology proposed by Lebart et al.*(41)* First, a principal component analysis of the data was conducted. Next, the number of clusters by a hierarchical cluster analysis was determined, and finally, a consolidation step by k-means clustering was performed.

After identification of those autoantibody-based subgroups, immunological characteristics were evaluated for each group. Cytokine concentrations were analyzed after log transformation. Linear regression models were fitted to estimate the differences in cytokines and lymphocyte populations among clusters. All models were adjusted by age and sex. Post-hoc comparison of means was based on both adjusted Bonferroni p-values and Fisher’s protected least significant differences procedure using t statistics based on Satterwhaite’s approximation. The significance level of the study was set to 0.05. Statistical analyses were done using R software version 4.0.2.

## Data Availability

All data are available in the main text or the supplementary materials.

## Supplementary Materials

Table S1. Cellular markers used for immunophenotyping of lymphocytes by flow cytometry.

Table S2. Differential expression of IgG autoantibodies.

Table S2. Differential expression of IgM autoantibodies.

## Acknowledgments

The authors would like to thank Yhojan Rodriguez, María Higuera, Daniela Polanía, and Mariana Borras for their assistance at the beginning of this work and all the members of the CREA for their contributions and fruitful discussions during the preparation of the manuscript.

## Author contributions

Conceptualization: JMA, GJT, CETG, RDM.

Acquisition of data: MRJ, EZ, ANP, ASA, CP, WZR, MAV, MR.

Methodology: CRS, YAA, DMM, LJRS, QZL, MR.

Statistical Analysis: MR, CZ.

Funding acquisition: CRS, JMA.

Project administration: CRS, JMA.

Supervision: JMA.

Writing – original draft: MR.

Writing – review & editing: MR, CRA, YAA, DMM.

## Data and materials availability

All data are available in the main text or the supplementary materials.

