## Supplementary Material 1 for "Polyautoimmunity Clusters as a New Taxonomy of Autoimmune Diseases"

This appendix has been provided by the authors to give readers additional information about their work.

**Most Recent Update:** August 10, 2021.

**Polyautoimmunity Clusters as a New Taxonomy of Autoimmune Diseases.**

**Supplementary Tables**

**Supplementary Table 1.** Cellular markers used for immunophenotyping of lymphocytes by flow cytometry.

| **Cell Subsets** | **Markers** |
| --- | --- |
| T cells | CD3^+^ |
| CD4+ T Cells | CD3^+^/CD4^+^ |
| Naïve CD4+ T cells | CD3^+^/CD4^+^/CD197^+^/CD45RO^-^ |
| Activated CD4+ T cells | CD3^+^/CD4^+^/CD38^+^/HLA-DR^+^ |
| Effector CD4+ T cells | CD3^+^/CD4^+^/CD197^-^/CD45RO^-^ |
| Effector memory CD4+ T cells | CD3^+^/CD4^+^/CD197^-^/CD45RO^+^ |
| Central memory CD4+ T cells | CD3^+^/CD4^+^/CD197^+^/CD45RO^+^ |
| CD8+ T Cells | CD3^+^/CD8^+^ |
| Naïve CD8+ T cells | CD3^+^/CD8^+^/CD197^+^/CD45RO^-^ |
| Activated CD8+ T cells | CD3^+^/CD8^+^/CD38^+^/HLA-DR^+^ |
| Effector CD8+ T cells | CD3^+^/CD8^+^/CD197^-^/CD45RO^-^ |
| Effector memory CD8+ T cells | CD3^+^/CD8^+^/CD197^-^/CD45RO^+^ |
| Central memory CD8+ T cells | CD3^+^/CD8^+^/CD197^+^/CD45RO^+^ |
| CD4+CD8+ T Cells | CD3^+^/CD4^+^/CD8^+^ |
| Th1 cells | CD3^+^/CD4^+^/CD194^-^/CD183^+^/CCR10^-^/CD196^-^ |
| Th2 cells | CD3^+^/CD4^+^/CD194^+^/CD183^-^/CCR10^-^/CD196^-^ |
| Th9 cells | CD3^+^/CD4^+^/ CD194^-^/CD196^+^ |
| Th17 cells | CD3^+^/CD4^+^/CD194^+^/CD183^-^/CCR10^-^/CD196^+^ |
| Th22 cells | CD3^+^/CD4^+^/CD194^+^/CD183^-^/CCR10^+^/CD196^+^ |
| Tregs | CD3^+^/CD4^+^/CD25^+^/CD127^low^ |
| Th17/Th1 cells | CD3^+^/CD4^+^/CD194^-^/CD183^+^/CCR10^-^/CD196^+^ |
| CD19+ CD20- B cells | CD19^+^/CD20^-^ |
| CD19+ CD20+ B cells | CD19^+^/CD20^+^ |
| Naïve B cells | CD19^+^/CD27^-^/IgD^+^ |
| Memory B cells | CD19^+^/CD27^+^/IgD^-^ |
| Non-classical memory B cells | CD19^+^/CD27^+^/IgD^+^ |
| Plasmablasts | CD19^+^/CD20^-^/CD27^+^/IgD^-^/CD24^-^/CD38^+^ |
| Transitional B cells | CD19^+^/CD24^High^/CD38^High^ |

Abbreviations: CD: Cluster of differentiation; Ig: Immunoglobulin; HLA: Human leukocyte antigens; Th: T helper; Tregs: Regulatory T cells.

**Supplementary Table 2.** Differential expression of IgG autoantibodies.

| Antigen | Log_2_ Fold change RA vs Healthy controls (Adjusted P value) | Log_2_ Fold change SLE vs Healthy controls (Adjusted P value) | Log_2_ Fold change SS vs Healthy controls (Adjusted P value) | Log_2_ Fold change AITD vs Healthy controls (Adjusted P value) | Log_2_ Fold change SSc vs Healthy controls (Adjusted P value) |
| --- | --- | --- | --- | --- | --- |
| Aggrecan | -0.26 (0.7673) | 0.93 (0.0678) | 0.81 (0.3716) | 1.70 (0.1130) | 1.34 (0.0937) |
| AGTR | -0.22 (0.9138) | -0.15 (0.8493) | 0.17 (0.7445) | -0.57 (0.2629) | 0.15 (0.8648) |
| Alpha Fodrin | -0.13 (0.8026) | 0.22 (0.7402) | 0.04 (0.7445) | -0.11 (0.7789) | 1.16 (0.8911) |
| Alpha-Actinine | 0.30 (0.7673) | 3.61 (0.1059) | 1.04 (0.9987) | 0.86 (0.5671) | 1.62 (0.4114) |
| AQP4 | -0.03 (0.9206) | 0.18 (0.4400) | 0.08 (0.8899) | 0.44 (0.2188) | 0.30 (0.4114) |
| B2 glycoprotein 1 | 0.23 (0.7673) | 0.13 (0.9333) | 0.36 (0.5389) | -0.26 (0.4012) | 0.09 (0.8795) |
| BPI | 0.20 (0.8698) | 0.33 (0.3140) | -0.17 (0.5935) | -0.68 (0.2107) | -0.65 (0.0898) |
| CENP-A | 0.86 (0.7673) | 1.89 (0.0740) | 0.42 (0.5095) | 0.68 (0.2107) | 6.71 (<0.0001) |
| CENP-B | 0.07 (0.9138) | 0.93 (0.1824) | 0.46 (0.5672) | 0.46 (0.3100) | 3.38 (<0.0001) |
| Chondroitin Sulfate C | 0.23 (0.9533) | 2.50 (0.0145) | 0.60 (0.5379) | -0.08 (0.2107) | 3.61 (0.0007) |
| Collagen I | 0.40 (0.7673) | 0.69 (0.0502) | 0.29 (0.5379) | -1.28 (0.2107) | -0.36 (0.8648) |
| Collagen II | -0.18 (0.7673) | 0.92 (0.0582) | -0.45 (0.8654) | -1.76 (0.7789) | -0.11 (0.3249) |
| Collagen III | 0.19 (0.7673) | 0.37 (0.0268) | 0.42 (0.3230) | -1.08 (0.3100) | 0.37 (0.3799) |
| Collagen IV | 0.11 (0.7673) | 0.38 (0.0051) | 0.35 (0.0202) | 0.19 (0.2107) | 0.27 (0.0966) |
| Collagen V | -0.04 (0.9018) | 0.41 (0.0636) | 0.27 (0.4685) | 0.17 (0.5105) | 0.43 (0.1911) |
| Collagen VI | -0.26 (0.7871) | 2.87 (<0.0001) | 0.46 (0.8158) | 1.09 (0.2107) | 1.14 (0.2836) |
| Complement C1q | -0.09 (0.7673) | 0.53 (0.1093) | 0.20 (0.6685) | 0.09 (0.9587) | 0.40 (0.3234) |
| Complement C3 | 0.13 (0.9919) | 0.26 (0.3710) | 0.00 (0.7330) | -0.45 (0.3186) | -0.10 (0.6598) |
| Complement C3a | 0.36 (0.7673) | 0.54 (0.0098) | 0.43 (0.1139) | -0.33 (0.5784) | 0.50 (0.0612) |
| Complement C4 | 0.03 (0.9533) | 0.20 (0.3557) | 0.08 (0.9591) | 0.29 (0.3100) | 0.29 (0.5551) |
| Complement C5 | 0.00 (0.9919) | 0.16 (0.1608) | 0.17 (0.4685) | -0.06 (0.8365) | 0.18 (0.4568) |
| Complement C6 | -0.09 (0.8344) | -0.01 (0.6658) | 0.08 (0.9908) | -0.15 (0.6520) | 0.05 (0.7255) |
| Complement C7 | 0.14 (0.7673) | 0.29 (0.0774) | 0.01 (0.9348) | -0.21 (0.7503) | -0.09 (0.6598) |
| Complement C8 | 0.00 (0.9919) | 0.24 (0.2634) | -0.16 (0.5095) | -0.36 (0.2432) | -0.02 (0.7416) |
| Complement C9 | 0.10 (0.7673) | 0.41 (0.0591) | 0.16 (0.5095) | -0.06 (0.8798) | 0.12 (0.6176) |
| Core Histone | 0.11 (0.9533) | 0.67 (0.0268) | 0.05 (0.7801) | -0.40 (0.4282) | 0.27 (0.7098) |
| CRP | -0.22 (0.7673) | 0.23 (0.0740) | 0.12 (0.5935) | -0.14 (0.6626) | 0.36 (0.1488) |
| Cytochrome C | 0.45 (0.9138) | 0.74 (0.0774) | 0.05 (0.8533) | -0.84 (0.9861) | 0.80 (0.1989) |
| DNA Polymerase beta | -0.15 (0.9533) | 0.17 (0.5463) | -0.14 (0.8654) | -0.65 (0.3599) | -0.30 (0.3234) |
| dsDNA | -0.35 (0.9533) | 4.39 (<0.0001) | 0.78 (0.5935) | 1.25 (0.2107) | 0.87 (0.8795) |
| EBNA1 | -0.10 (0.9999) | 0.50 (0.3067) | -0.27 (0.9908) | -0.22 (0.8365) | -0.88 (0.2490) |
| Elastin | 0.11 (0.8344) | 1.26 (0.0010) | 0.02 (0.9987) | -0.50 (0.9306) | 0.11 (0.8971) |
| Entaktin EDTA | .26 (0.7673) | 2.22 (0.0003) | 0.78 (0.2225) | 0.59 (0.2107) | 3.47 (<0.0001) |
| Factor B | -0.05 (0.9138) | 0.11 (0.2634) | -0.04 (0.8791) | -0.21 (0.9717) | 0.11 (0.4598) |
| Factor D | 0.74 (0.7673) | 2.04 (0.0651) | 0.38 (0.3889) | 0.74 (0.2059) | 0.91 (0.4114) |
| Factor H | -0.32 (0.7673) | -0.22 (0.7841) | -0.51 (0.5935) | 0.16 (0.8928) | -0.13 (0.7832) |
| Factor I | -0.21 (0.9261) | 0.21 (0.3357 | -0.31 (0.7603) | -0.46 (0.3546) | -0.36 (0.4598) |
| Fibrinogen IV | 0.16 (0.7673) | 0.88 (0.0651) | 0.18 (0.7330) | 0.01 (0.2933) | 0.33 (0.3234) |
| Fibrinogen S | 0.11 (0.7673) | 0.55 (0.1542) | 0.36 (0.4073) | 0.13 (0.3527) | 0.12 (0.4858) |
| Fibronectin | -0.72 (0.9533) | 0.29 (0.0440) | -0.02 (0.5935) | 0.26 (0.2395) | 0.42 (0.0431) |
| GBM | 0.54 (0.9138) | 1.20 (0.0802) | -0.12 (0.5732) | -0.16 (0.5211) | 0.70 (0.9196) |
| Genomic DNA | -0.20 (0.9533) | 6.62 (0.0005) | 2.44 (0.5095) | 2.75 (0.1484) | 3.52 (0.2490) |
| Gliadin | 0.25 (0.7673) | 0.54 (0.1771) | 0.68 (0.3716) | 0.73 (0.2107) | 0.51 (0.3779) |
| Glycated Albumin | -0.09 (0.8755) | 0.12 (0.2102) | 0.05 (0.9908) | -0.53 (0.2107) | 0.08 (0.8795) |
| GP2 | 0.07 (0.7673) | -0.04 (0.7841) | 0.40 (0.3716) | -0.58 (0.2107) | 0.18 (0.6176) |
| GP210 | 1.25 (0.7673) | 2.24 (0.0003) | 1.05 (0.3716) | -1.45 (0.2107) | 0.02 (0.8284) |
| Hemocyanin | 0.02 (0.7871) | 0.15 (0.1416) | 0.13 (0.5379) | 0.07 (0.4282) | 0.30 (0.0979) |
| Heparan sulfate proteoglycan | 0.13 (0.7673) | 1.15 (0.1059) | 0.72 (0.3716) | -1.13 (0.5784) | 1.12 (0.2456) |
| Heparan Sulphate | 0.25 (0.7673) | 0.59 (0.0184) | 0.09 (0.3716) | -2.89 (0.2795) | -0.35 (0.5374) |
| Heparin | 0.94 (0.8026) | 3.51 (0.0001) | 0.69 (0.5935) | -2.18 (0.7789) | -0.75 (0.7083) |
| Histone H1 | 0.04 (0.7673) | 1.36 (0.0125) | -0.18 (0.6644) | 0.02 (0.6185) | -0.38 (0.4424) |
| Histone H2A | -0.02 (0.9138) | 1.22 (0.0047) | 0.03 (0.7981) | 0.07 (0.9861) | 0.07 (0.9188) |
| Histone H2B | -0.06 (0.9533) | 0.87 (0.0051) | 0.10 (0.3989) | -0.07 (0.5671) | 0.05 (0.6971) |
| Histone H3 | 0.20 (0.7673) | 0.45 (0.0502) | 0.16 (0.3716) | 0.12 (0.9468) | 0.57 (0.2358) |
| Histone H4 | 0.02 (0.7871) | 0.58 (0.0440) | 0.36 (0.1382) | 0.37 (0.2107) | 1.10 (0.0003) |
| Intrinsic Factor | -0.28 (0.9533) | -0.21 (0.7901) | 0.12 (0.7214) | -0.45 (0.9717) | -0.09 (0.7255) |
| Jo-1 | -0.02 (0.8698) | 0.16 (0.4373) | -0.10 (0.5379) | -0.26 (0.6520) | -0.07 (0.6598) |
| KU (P70/P80) | 0.10 (0.8026) | 0.48 (0.0440) | 0.11 (0.5672) | -0.20 (0.4507) | 0.23 (0.3234) |
| La/SSB | 0.11 (0.7673) | 0.48 (0.1542) | 0.85 (0.2225) | 0.34 (0.2458) | 0.49 (0.1261) |
| Laminin | 0.23 (0.7673) | 1.67 (0.0054) | 1.94 (0.0012) | -1.06 (0.8648) | 0.92 (0.2490) |
| LC1 | -1.58 (0.7673) | -0.19 (0.3976) | 2.27 (0.3716) | 5.04 (0.2107) | 6.70 (0.0132) |
| LKM1 | 0.15 (0.7673) | -0.30 (0.2996) | -0.92 (0.8791) | -1.40 (0.3954) | 0.73 (0.0328) |
| LPS | .06 (0.9533) | 0.83 (0.0582) | 0.13 (0.9794) | 0.36 (0.6520) | 0.31 (0.8665) |
| M2 | 0.02 (0.8755) | 0.75 (0.0802) | 0.22 (0.3947) | -0.45 (0.3320) | 0.33 (0.2490) |
| Matrigel | 0.08 (0.7673) | 1.18 (0.0582) | 0.54 (0.3716) | -1.34 (0.5694) | 0.69 (0.3234) |
| MDA5 | 0.17 (0.9076) | 1.05 (0.1096) | 0.30 (0.9987) | 0.42 (0.9468) | 0.66 (0.7416) |
| Mi-2 | 0.05 (0.7673) | 0.55 (0.0213) | -0.31 (0.6753) | -1.06 (0.9861) | 0.41 (0.1252) |
| Mitochondrial antigen | -0.03 (0.9138) | 0.96 (0.1059) | 0.64 (0.3716) | 0.09 (0.3037) | 0.99 (0.2358) |
| MPO | -0.01 (0.9138) | 0.28 (0.0904) | 0.23 (0.3887) | -0.33 (0.2176) | 0.50 (0.0431) |
| Muscarinic receptor | -0.45 (0.9500) | 0.30 (0.4373) | 0.42 (0.5209) | 0.36 (0.6993) | 0.51 (0.2490) |
| Myelin basic protein | 0.03 (0.7673) | 0.19 (0.7841) | -0.22 (0.5672) | -0.61 (0.5041) | 0.17 (0.9188) |
| Myosin | 0.29 (0.7673) | 0.80 (0.0010) | 0.33 (0.3716) | -0.48 (0.2176) | 0.45 (0.1405) |
| Nucleolin | 0.20 (0.9533) | 0.46 (0.0678) | 0.08 (0.4685) | -1.62 (0.2107) | 0.30 (0.8795) |
| Nucleosome antigen | -0.01 (0.9533) | 0.89 (0.0093) | -0.02 (0.9591) | 0.43 (0.3186) | 0.28 (0.8648) |
| Nup 62 | 0.65 (0.7673) | 0.18 (0.5513) | -0.64 (0.7620) | -2.18 (0.9717) | -1.64 (0.2490) |
| PCNA | 0.10 (0.7673) | 0.56 (0.0184) | 0.08 (0.9656) | -0.13 (0.5671) | 0.27 (0.3234) |
| Peroxiredoxin 1 | 0.17 (0.7673) | 0.24 (0.0904) | 0.21 (0.5095) | -0.06 (0.7829) | 0.24 (0.4598) |
| PL-12 | 0.21 (0.7673) | 0.48 (0.0774) | -0.27 (0.9656) | -0.74 (0.1130) | 0.06 (0.8665) |
| PL-7 | 0.03 (0.9533) | 0.21 (0.2113) | -0.24 (0.3716) | -0.58 (0.2107) | 0.10 (0.7407) |
| PM/Scl 100 | -0.14 (0.7871) | 0.88 (0.0824) | 0.77 (0.3716) | -0.08 (0.7241) | 1.08 (0.2475) |
| PM/Scl-75 | 0.01 (0.9138) | 0.32 (0.2377) | 0.13 (0.9348) | -0.19 (0.6520) | 0.46 (0.4598) |
| PR3 | -0.62 (0.8344) | 0.21 (0.0440) | 0.29 (0.3758) | -1.49 (0.2395) | 0.57 (0.0439) |
| Proteoglycan | 0.91 (0.7673) | 1.02 (0.1041) | 0.21 (0.6685) | -2.11 (0.4282) | -1.61 (0.3234) |
| Prothrombin protein | -0.01 (0.9533) | 0.28 (0.2267) | 0.03 (0.9987) | -0.40 (0.4046) | 0.01 (0.9122) |
| Ribo Phosphoprotein P0 | 0.13 (0.7673) | 1.99 (0.0042) | 0.53 (0.1319) | 0.30 (0.2176) | 1.26 (0.1261) |
| Ribo Phosphoprotein P1 | 0.15 (0.7673) | 1.99 (0.0440) | 1.01 (0.0040) | 0.22 (0.2107) | 1.30 (0.0443) |
| Ribo Phosphoprotein P2 | -0.06 (0.9999) | 1.52 (0.0085) | 0.15 (0.9656) | 0.32 (0.6520) | 0.60 (0.2490) |
| Ro/SSA (52 Kda) | 0.58 (0.7673) | 1.53 (0.0023) | 2.78 (< 0.0001) | 0.73 (0.1484) | 1.89 (0.0014) |
| Ro/SSA (60 Kda) | 0.15 (0.7673) | 1.19 (0.0356) | 1.66 (0.0158) | 0.04 (0.2320) | 0.28 (0.2055) |
| S100 | -0.03 (0.9138) | 0.38 (0.3067) | -0.07 (0.5732) | 0.00 (0.7829) | -0.28 (0.4598) |
| Scl-70/Topoisomerase I | 0.23 (0.8344) | 0.66 (0.0582) | 0.53 (0.3989) | -0.15 (0.9348) | 0.18 (0.4424) |
| Sm | -0.13 (0.7871) | 1.69 (0.1059) | 0.54 (0.2225) | -0.77 (0.6939) | 0.92 (0.0452) |
| Sm/RNP | 0.01 (0.8344) | 2.80 (0.0023) | 0.66 (0.2225) | 0.59 (0.2107) | 0.74 (0.2490) |
| SmD | -0.01 (0.8103) | 1.68 (0.0740) | 0.33 (0.5095) | -0.85 (0.3599) | 0.80 (0.0472) |
| SmD1 | 0.25 (0.7673) | 1.80 (0.0219) | 0.40 (0.5095) | -0.25 (0.7829) | 0.02 (0.8648) |
| SmD2 | -0.29 (0.7673) | 0.54 (0.2798) | -0.10 (0.5379) | -1.57 (0.2176) | -0.01 (0.4598) |
| SmD3 | 0.47 (0.7673) | 1.38 (0.1286) | 0.01 (0.9591) | -2.33 (0.2107) | 0.62 (0.3234) |
| SRP54 | -0.87 (0.7673) | -0.09 (0.6897) | -0.16 (0.7445) | 0.29 (0.9540) | -0.10 (0.9188) |
| ssDNA | -0.24 (0.9325) | 2.35 (<0.0001) | 0.67 (0.3716) | 1.28 (0.1130) | 1.13 (0.0431) |
| ssRNA | 0.88 (0.8026) | 6.93 (0.0005) | 0.60 (0.4685) | 0.83 (0.2107) | -0.77 (0.5433) |
| T1F1 gamma | 0.18 (0.7673) | 1.08 (0.0774) | 0.47 (0.4685) | 0.17 (0.5461) | 0.67 (0.0144) |
| Thyroglobulin | 1.13 (0.7673) | 1.36 (0.0356) | 1.74 (0.0061) | 3.27 (<0.0001) | 1.62 (0.0144) |
| TNF | -0.01 (0.9018) | 0.01 (0.7841) | 0.20 (0.7603) | -0.02 (0.7275) | -0.24 (0.3570) |
| TPO | 0.41 (0.7673) | 0.09 (0.5155) | 0.38 (0.3758) | 1.40 (0.0022) | -0.23 (0.6598) |
| TTG | 0.08 (0.7871) | 0.38 (0.0664) | 0.16 (0.5379) | -0.24 (0.3173) | -0.12 (0.4598) |
| U1-snRNP 68/70 | 0.35 (0.7673) | 1.75 (0.0055) | 0.92 (0.0802) | 0.59 (0.2107) | 1.21 (0.1252) |
| U1-snRNP A | 0.29 (0.7673) | 1.25 (0.0219) | 0.33 (0.3716) | 0.44 (0.2107) | 0.25 (0.3234) |
| U1-snRNP B/B' | 0.68 (0.7673) | 2.85 (0.0010) | 3.38 (0.0003) | 1.11 (0.1484) | 2.56 (<0.0001) |
| U1-snRNP C | 0.49 (0.7673) | 2.02 (0.0028) | 2.12 (0.0025) | 1.42 (0.1484) | 2.29 (0.0003) |
| Vimentin | 0.25 (0.7673) | 0.47 (0.0356) | 0.14 (0.2225) | -0.77 (0.2529) | 0.78 (0.0966) |
| Vitronectin | 0.27 (0.7673) | 0.50 (0.0664) | 0.24 (0.7214) | -0.28 (0.4282) | 0.11 (0.9601) |

^a^ Data was analyzed by *t*-test, and adjusted p values were obtained by false discovery rate. RA: Rheumatoid arthritis; SLE: Systemic lupus erythematosus; SS: Sjögren’s syndrome; AITD: Autoimmune thyroid disease; SSc: Systemic sclerosis.

**Supplementary Table 3.** Differential expression of IgM autoantibodies.

| Antigen | Log_2_ Fold change RA vs Healthy controls (Adjusted P value) | Log_2_ Fold change SLE vs Healthy controls (Adjusted P value) | Log_2_ Fold change SS vs Healthy controls (Adjusted P value) | Log_2_ Fold change AITD vs Healthy controls (Adjusted P value) | Log_2_ Fold change SSc vs Healthy controls (Adjusted P value) |
| --- | --- | --- | --- | --- | --- |
| Aggrecan | -0.50 (0.7222) | -0.67 (0.6749) | 0.37 (0.6962) | 1.72 (0.3461) | 2.15 (0.0546) |
| AGTR | 0.08 (0.9813) | -0.32 (0.3166) | 0.14 (0.9607) | 0.06 (0.9967) | 0.18 (0.7534) |
| Alpha Fodrin | 0.16 (0.9813) | -0.22 (0.4113) | 0.22 (0.9064) | 0.44 (0.5625) | 0.48 (0.2887) |
| AQP4 | 0.40 (0.9813) | -0.08 (0.2987) | 0.39 (0.9950) | 0.60 (0.9350) | 0.38 (0.9618) |
| B2 glycoprotein 1 | 0.18 (0.9813) | -0.30 (0.2987) | 0.17 (0.9950) | -0.10 (0.9942) | 0.02 (0.7534) |
| BPI | 0.10 (0.9813) | -0.39 (0.2987) | -0.09 (0.9950 | -0.32 (0.8937) | -0.42 (0.6212) |
| CENP-A | 0.22 (0.9813) | -0.05 (0.7311) | 0.02 (0.9950) | 0.00 (0.9967) | 0.86 (0.0159) |
| CENP-B | -0.01 (0.9813) | -0.39 (0.2987) | -0.07 (0.9950) | 0.07 (0.9350) | 0.64 (0.0370) |
| Collagen V | 0.18 (0.9813) | -0.25 (0.3407) | 0.44 (0.9950) | 0.61 (0.9350) | 0.55 (0.7534) |
| Collagen VI | 0.15 (0.9813) | 0.90 (0.0020) | 0.51 (0.2073) | 0.51 (0.1830) | 0.19 (0.6303) |
| Complement C1q | -0.07 (0.9813) | -0.41 (0.2987) | 0.07 (0.9950) | 0.30 (0.5879) | 0.23 (0.7534) |
| Complement C3 | 0.13 (0.9813) | -0.38 (0.2987) | -0.09 (0.9950) | -0.28 (0.9350) | -0.32 (0.7534) |
| Complement C3a | 0.21 (0.9813) | -0.08 (0.9433) | -0.31 (0.6962) | -0.16 (0.9711) | -0.71 (0.2368) |
| Complement C4 | 0.01 (0.9813) | -0.36 (0.2987) | 0.04 (0.9950) | -0.03 (0.9967) | 0.01 (0.9929) |
| Complement C5 | -0.07 (0.9813) | -0.50 (0.2987) | -0.07 (0.9950) | -0.12 (0.9942) | -0.02 (0.9929) |
| Complement C6 | 0.08 (0.9813) | -0.36 (0.2987) | 0.05 (0.9950) | -0.04 (0.9967) | -0.02 (0.9929) |
| Complement C7 | 0.06 (0.9813) | -0.40 (0.2987) | -0.02 (0.9950) | -0.26 (0.9350) | -0.23 (0.9929) |
| Complement C8 | 0.02 (0.9813) | -0.39 (0.2987) | 0.06 (0.9950) | -0.1 (0.9967) | -0.09 (0.9929) |
| Complement C9 | -0.06 (0.9813) | -0.56 (0.2987) | -0.1 (0.9950) | -0.14 (0.9711) | -0.08 (0.9929) |
| Core Histone | 0.14 (0.9813) | -0.76 (0.3407) | -1.09 (0.5638) | -0.93 (0.5879) | -1.18 (0.0421) |
| CRP | -0.08 (0.9813) | -0.48 (0.2987) | 0.05 (0.9950) | 0.15 (0.9350 | -0.02 (0.9929) |
| Cytochrome C | 0.33 (0.9813) | -0.05 (0.2987) | 0.63 (0.9950) | -0.11 (0.9350) | -0.27 (0.9929) |
| DNA Polymerase beta | -0.09 (0.9813) | -0.34 (0.2987) | 0.06 (0.9950) | 0.16 (0.9350) | 0.12 (0.9618) |
| dsDNA | 0.23 (0.9813) | 1.93 (0.0019) | 0.88 (0.0799) | 0.89 (0.0889) | 0.34 (0.4085) |
| Elastin | 0.41 (0.9813) | 0.10 (0.6762) | 0.21 (0.9950) | -0.14 (0.9350) | -0.54 (0.7534) |
| Entaktin EDTA | 0.20 (0.9813) | -0.08 (0.9001) | -0.06 (0.9950) | -0.14 (0.9350) | -0.3 (0.4085) |
| Factor B | 0.06 (0.9813) | -0.41 (0.2987) | -0.05 (0.9950) | -0.2 (0.9350) | -0.19 (0.9929) |
| Factor D | 0.61 (0.9813) | 0.35 (0.9629) | 0.32 (0.9950) | -0.29 (0.9350) | -0.29 (0.7534) |
| Factor H | 0.15 (0.9813) | -0.29 (0.2987) | -0.01 (0.9950) | -0.20 (0.9350) | -0.26 (0.9160) |
| Factor I | 0.08 (0.9813) | -0.37 (0.2987) | -0.05 (0.9950) | -0.26 (0.9350) | -0.18 (0.9929) |
| Fibrinogen IV | 0.08 (0.9813) | -0.20 (0.3500) | 0.15 (0.9950) | -0.17 (0.9967) | 0.02 (0.7534) |
| Fibrinogen S | 0.07 (0.9813) | -0.26 (0.3376) | 0.16 (0.9950) | -0.08 (0.9967) | 0.04 (0.7534) |
| Fibronectin | 0.09 (0.9813) | 0.56 (0.9001) | 0.51 (0.9064) | 0.55 (0.5879) | 1.19 (0.1579) |
| GBM | 0.03 (0.9813) | -0.28 (0.3069) | 0.07 (0.9950) | -0.20 (0.9967) | -0.23 (0.9929) |
| Gliadin | 0.12 (0.9813) | -0.29 (0.2987) | 0.07 (0.9950) | 0.13 (0.9350) | -0.06 (0.9929) |
| Glycated Albumin | -0.07 (0.9813) | -0.55 (0.0981) | -0.16 (0.9950) | -0.17 (0.9350) | -0.17 (0.9929) |
| GP2 | -0.01 (0.9813) | -0.41 (0.2987) | 0.01 (0.9950) | -0.04 (0.9967) | -0.09 (0.9929) |
| GP210 | 0.27 (0.9813) | -0.16 (0.6769) | 0.07 (0.9950) | -0.14 (0.9711) | -0.51 (0.9929) |
| Hemocyanin | -0.13 (0.9813) | -0.51 (0.2987) | -0.03 (0.9950) | 0.03 (0.9407) | 0.03 (0.9929) |
| Heparan sulfate proteoglycan | 0.42 (0.9813) | 0.12 (0.7311) | 0.51 (0.5638) | -0.28 (0.9711) | -0.18 (0.9929) |
| Histone H1 | -0.48 (0.9813) | -1.10 (0.4217) | -2.77 (0.5638) | -2.90 (0.5625) | -3.98 (0.0284) |
| Histone H2A | -0.15 (0.9813) | -0.68 (0.4248) | -2.01 (0.4154) | -1.32 (0.4111) | -2.81 (0.0284) |
| Histone H2B | 0.12 (0.9813) | -0.45 (0.7267) | -1.04 (0.5638) | -0.37 (0.5879) | -1.62 (0.0370) |
| Histone H3 | 0.16 (0.9813) | -0.24 (0.3255) | -0.02 (0.9950) | -0.04 (0.9350) | -0.32 (0.5657) |
| Histone H4 | -0.15 (0.9813) | -0.39 (0.2987) | 0.03 (0.9950) | 0.45 (0.4111) | 0.23 (0.7534) |
| Insulin | 0.99 (0.7222) | 0.29 (0.9001) | 0.34 (0.9950) | -1.12 (0.3811) | -0.79 (0.7534) |
| Intrinsic Factor | 0.16 (0.9813) | -0.32 (0.3069) | 0.06 (0.9950) | -0.29 (0.9350) | -0.23 (0.7534) |
| Jo-1 | 0.04 (0.9813) | -0.44 (0.2987) | -0.12 (0.9950) | -0.22 (0.9350) | -0.29 (0.7534) |
| KU (P70/P80) | -0.02 (0.9813) | -0.39 (0.3407) | 0.03 (0.9950) | 0.15 (0.9350) | 0.15 (0.8457) |
| La/SSB | -0.05 (0.9813) | -0.35 (0.6060) | 0.00 (0.9950) | -0.21 (0.9350) | -0.16 (0.9545) |
| Laminin | 0.19 (0.9813) | 0.05 (0.6549) | 0.53 (0.2245) | 0.08 (0.8206) | 0.00 (0.7534) |
| LC1 | -1.63 (0.7222) | -0.63 (0.7271) | 2.21 (0.5638) | 4.89 (0.1830) | 6.12 (0.0313) |
| LKM1 | 0.06 (0.9813) | -1.48 (0.0981) | -1.44 (0.5638) | -1.15 (0.5879) | -1.67 (0.1704) |
| LPS | 0.33 (0.9813) | 0.87 (0.2272) | 0.20 (0.9064) | 0.45 (0.5625) | -0.32 (0.6277) |
| M2 | -0.1 (0.9813) | -0.40 (0.2987) | -0.12 (0.995) | -0.35 (0.8937) | -0.29 (0.9618) |
| Matrigel | 0.22 (0.9813) | -0.04 (0.9001) | 0.26 (0.7497) | -0.11 (0.9942) | -0.12 (0.9545) |
| MDA5 | -0.07 (0.9813) | -0.34 (0.2987) | 0.04 (0.995) | 0.18 (0.9350) | 0.10 (0.9618) |
| Mitochondrial antigen | 0.08 (0.9813) | -0.16 (0.4266) | 0.22 (0.6836) | 0.18 (0.7540) | 0.28 (0.4374) |
| MPO | 0.10 (0.9813) | -0.20 (0.6549) | -0.37 (0.4154) | -0.65 (0.1830) | -0.71 (0.0313) |
| Muscarinic receptor | -0.05 (0.9813) | -0.37 (0.2987) | -0.04 (0.9950) | 0.15 (0.9350) | 0.14 (0.7534) |
| Myelin basic protein | -0.42 (0.9813) | -0.78 (0.9629) | -2.37 (0.4154) | -1.66 (0.5625) | -2.87 (0.0284) |
| Myosin | 0.06 (0.9813) | -0.31 (0.3928) | -0.02 (0.9950) | -0.11 (0.9777) | -0.16 (0.9545) |
| Nucleolin | 0.04 (0.9813) | -0.38 (0.2987) | 0.06 (0.9950) | -0.24 (0.935) | -0.21 (0.9929) |
| Nucleosome antigen | 0.05 (0.9813) | 0.05 (0.8201) | -1.41 (0.9064) | -0.43 (0.8937) | -1.32 (0.1704) |
| PCNA | -0.01 (0.9813) | -0.46 (0.2987) | -0.07 (0.9950) | -0.03 (0.9967) | -0.16 (0.7534) |
| Peroxiredoxin 1 | 0.05 (0.9813) | -0.33 (0.2987) | 0.08 (0.9950) | -0.10 (0.9967) | 0.03 (0.9705) |
| PL-12 | 0.04 (0.9813) | -0.36 (0.2987) | 0.01 (0.9950) | -0.22 (0.9350) | -0.14 (0.9929) |
| PL-7 | 0.05 (0.9813) | -0.43 (0.2987) | -0.16 (0.9950) | -0.28 (0.8937) | -0.35 (0.6623) |
| PM/Scl 100 | 0.00 (0.9813) | -0.33 (0.2987) | -0.01 (0.9950) | -0.05 (0.9967) | -0.03 (0.9929) |
| PM/Scl-75 | 0.03 (0.9813) | -0.38 (0.2987) | 0.09 (0.9950) | 0.11 (0.9350) | 0.10 (0.9007) |
| Prothrombin protein | 0.12 (0.9813) | -0.23 (0.3407) | -0.03 (0.9950) | -0.12 (0.9967) | -0.09 (0.9929) |
| Ribo Phosphoprotein P0 | -0.05 (0.9813) | -0.30 (0.5999) | -0.05 (0.9950) | 0.14 (0.9350) | 0.04 (0.9929) |
| Ribo Phosphoprotein P1 | -0.06 (0.9813) | -0.06 (0.9001) | 0.11 (0.9950) | 0.37 (0.5879) | 0.31 (0.6644) |
| Ribo Phosphoprotein P2 | 0.44 (0.9813) | 0.69 (0.2987) | 0.40 (0.9950) | 0.33 (0.9350) | 0.10 (0.5569) |
| Ro/SSA (52 Kda) | 0.20 (0.9813) | 0.00 (0.9001) | 0.93 (0.0799) | 0.45 (0.4111) | 0.23 (0.6623) |
| Ro/SSA (60 Kda) | 0.11 (0.9813) | -0.23 (0.3760) | 0.05 (0.9950) | -0.23 (0.9350) | -0.34 (0.7534) |
| S100 | 0.06 (0.9813) | -0.39 (0.2987) | -0.21 (0.7658) | -0.27 (0.8937) | -0.55 (0.2860) |
| Scl-70/Topoisomerase I | 0.14 (0.9813) | -0.33 (0.2987) | -0.12 (0.9950) | -0.4 (0.7540) | -0.41 (0.6623) |
| Sm | 0.14 (0.9813) | 0.06 (0.6549) | 0.08 (0.9950) | -0.1 (0.9350) | -0.33 (0.7534) |
| Sm/RNP | 0.01 (0.9813) | 0.19 (0.2987) | -0.02 (0.9950) | 0.08 (0.9350) | 0.06 (0.9160) |
| SmD | 0.00 (0.9813) | -0.13 (0.9001) | -0.40 (0.5638) | -0.12 (0.7630) | -0.37 (0.1579) |
| SmD1 | 0.18 (0.9813) | 0.05 (0.8201) | -0.43 (0.5638) | -0.66 (0.5625) | -1.17 (0.0421) |
| SmD2 | -0.09 (0.9813) | -0.31 (0.606) | -0.59 (0.5638) | -0.57 (0.5879) | -0.96 (0.0900) |
| SmD3 | 0.33 (0.9813) | 0.03 (0.9001) | -0.34 (0.5638) | -0.7 (0.5625) | -0.99 (0.0546) |
| SRP54 | -0.15 (0.9813) | -0.31 (0.3389) | 0.20 (0.6962) | 0.57 (0.2299) | 0.66 (0.0546) |
| ssDNA | 0.42 (0.7222) | 1.92 (<0.001) | 0.97 (0.0536) | 1.28 (0.0037) | 0.81 (0.0546) |
| T1F1 gama | 0.01 (0.9813) | -0.39 (0.4643) | -0.15 (0.9950) | -0.04 (0.9350) | -0.34 (0.2368) |
| Thyroglobulin | 1.09 (0.9813) | 0.84 (0.9001) | 0.96 (0.9950) | 1.54 (0.8206) | 1.28 (0.9929) |
| TNF | 0.08 (0.9813) | -0.46 (0.2987) | -0.1 (0.9950) | -0.15 (0.9350) | -0.30 (0.7534) |
| TPO | 0.13 (0.9813) | -0.28 (0.2987) | 0.08 (0.9950) | 0.09 (0.9350) | -0.05 (0.9929) |
| TTG | 0.00 (0.9813) | -0.40 (0.2987) | -0.1 (0.9950) | -0.28 (0.8937) | -0.27 (0.7534) |
| U1-snRNP 68/70 | 0.38 (0.9813) | 0.04 (0.9001) | -0.35 (0.5638) | -0.64 (0.5625) | -0.88 (0.0672) |
| U1-snRNP A | 0.07 (0.9813) | -0.09 (0.9001) | 0.11 (0.9950) | 0.22 (0.7216) | 0.07 (0.9160) |
| U1-snRNP B/B' | -0.31 (0.9813) | -0.40 (0.4116) | -1.4 (0.4154) | -0.44 (0.5625) | -1.7 (0.0284) |
| U1-snRNP C | 0.13 (0.9813) | -0.26 (0.3407) | -0.34 (0.7497) | 0.21 (0.9967) | 0.04 (0.9929) |
| Vimentin | 0.49 (0.9813) | 0.39 (0.6769) | 0.88 (0.3897) | 0.56 (0.5879) | 0.59 (0.4085) |
| Vitronectin | 0.23 (0.9813) | -0.24 (0.3389) | 0.11 (0.9950) | 0.06 (0.9350) | -0.08 (0.9929) |

^a^ Data was analyzed by *t*-test, and adjusted p values were obtained by false discovery rate. RA: Rheumatoid arthritis; SLE: Systemic lupus erythematosus; SS: Sjögren’s syndrome; AITD: Autoimmune thyroid disease; SSc: Systemic sclerosis.
